# Going beyond “regular and casual”: development of a classification of sexual partner types to enhance partner notification for sexually transmitted infections, a mixed methods approach

**DOI:** 10.1101/2020.09.27.20202705

**Authors:** Claudia Estcourt, Paul Flowers, Jackie Cassell, Maria Pothoulaki, Gabriele Vojt, Fiona Mapp, Melvina Woode Owusu, Nicola Low, John Saunders, Merle Symonds, Alison Howarth, Sonali Wayal, Rak Nandwani, Susie Brice, Alex Comer, Anne M Johnson, Catherine Mercer

## Abstract

**Objectives:** To develop a classification of sexual partner types for use in partner notification (PN) and other interventions to prevent sexually transmitted infections (STI).

**Methods:** A four-step process: 1) an iterative synthesis of five sources of evidence: scoping review of social and health sciences literature on partner types; analysis of relationship types in dating apps; systematic review of PN intervention content; review of PN guidelines; qualitative interviews with public, patients and health professionals, to generate an initial comprehensive classification; 2) multidisciplinary clinical expert consultation to revise the classification; 3) piloting of the revised classification in sexual health clinics during a randomised controlled trial of PN; 4) application of the Theoretical Domains Framework (TDF) to identify index patients’ willingness to engage in PN for each partner type.

**Results:** Five main partner types emerged from the evidence synthesis and consultation: ‘Established partner’, ‘New partner’, ‘Occasional partner’, ‘One-off partner’ and ‘Sex worker’. The types differed across several dimensions, including likely perceptions of sexual exclusivity, likelihood of sex reoccurring between index patient and sex partner. Sexual health professionals found the classification easy to operationalise. During the trial, they assigned all 3288 partners described by 2223 index patients to a category. The TDF analysis suggested that the partner types might be associated with different risks of STI reinfection, onward transmission and index patients’ engagement with PN.

**Discussion:** We developed an evidence-informed, useable classification of five sexual partner types to underpin PN practice and other STI prevention interventions. Analysis of biomedical, psychological and social factors that distinguish different partner types shows how each could warrant a tailored PN approach. This classification could facilitate the use of partner-centred outcomes. Additional studies are needed to determine the utility of the classification to improve measurement of the impact of PN strategies and help focus resources.

**Key messages:**

1. Current classifications of sexual partners limit understanding of STI transmission dynamics and hinder targeting and tailoring of partner notification interventions.
2. The limits and constraints of current classifications, together with recent socio-sexual changes, mean that a new classification is needed.
3. We developed a comprehensive, evidence-based classification of sexual partner types for use in partner notification that characterised and distinguished between partner and partnership types.
4. The five partner categories were readily adopted and easily operationalised in UK sexual health services.

## Introduction

Understanding the nature and number of sexual partners of people with sexually transmitted infections (STIs) is fundamental to understanding the epidemiology of STIs, delivery of high quality clinical care, and prevention of transmission through effective partner notification (PN).^1–6^ However, we need appropriate tools to assess to whom and how interventions should be targeted.

Specialist sexual and public health guidance and published researchers tend to use a simple dichotomy of “regular” (“steady”) or “casual” sexual partners.^3,7–10^ These categories do not take into account the complexities of sexual partnerships in ways that help understand the potential for STI transmission, or support clinical, research or prevention practice. The outcomes of PN generally specify an overall number of partners contacted/treated per index patient,^3^ ignoring variation in the timing and types of partnerships, the likelihood of onward transmission by partnership type,^1^ or the different kinds of support needed to notify partners effectively.^6^

The way people meet sex partners is changing. Through increasing internet use,^11^ online commercial socio/sexual networking sites have generated their own partner classifications, shaping the ways people understand and talk about relationships.^12^ Public awareness of different types of sexual partners is also increasing, with recognition of sexual interactions where the label of ‘partner’ is not applicable. These changes are taking place at a time of sustained rises in STIs in some groups.^13^

Social epidemiologists and behavioural scientists have sought to develop alternative ways of classifying partnership type to try and better understand STI and HIV risk (e.g.^14–16^), but there is no consensus. As a result, we lack comparable quantitative data for epidemiological studies and service evaluations. A standardised partner type classification, with face validity for both patients and service providers would improve measurement of the impact of PN strategies and help focus resources.^17^ If applied to the practice of PN, a new classification would help a move towards partner-centred outcomes (e.g. transmissions prevented according to partnership type) rather than patient-centred outcomes (e.g. partners tested/treated per index case).

The objectives of this study were to create a useable classification of sexual partners to improve the targeting of PN and other STI prevention interventions. The study addressed four questions: 1) Which labels are used to classify sexual partners and which biomedical, psychological and social aspects differentiate them? 2) What does a classification of sexual partners look like for clinical practice? 3) Is this classification acceptable and feasible for routine use? 4) How could use of the classification help to improve individual and public health?

## Methods

This study is part of the Limiting Undetected Sexually Transmitted infections to RedUce Morbidity (LUSTRUM; lustrum.org.uk) research programme. We used mixed research methods within a broad, biopsychosocial approach, acknowledging the importance of psychological, social and cultural factors to the understanding of sexual partnerships.^18^ We used a four-step process: 1) integrating evidence from diverse sources to develop an initial classification, 2) multidisciplinary clinical expert input to revise the classification, 3) piloting the classification in sexual health clinics, 4) application of the Theoretical Domains Framework (TDF)^19^ to explore the need for tailored PN.

1. **Evidence integration:** We iteratively synthesised the findings from four diverse sources of evidence: i) a scoping review of partnership types described in the social and health sciences literature,^20^ ii) a review of PN guidelines,^21^ iii) focus group discussions with lay people, including those recently diagnosed with an STI,^22^ and iv) a review of partnership types described in dating apps.^12^ The methods used for each source have been published separately. ^12,20–22^ We created a matrix of partner types, according to the key biomedical, psychological and social factors that differentiated them. First, we derived descriptions of partner types from the review of social and health sciences literature.^20^ Second, we used constant comparative techniques, i.e., taking published data and comparing them with our emerging findings and putting ‘like with like’, to map descriptions of types of relationship and concomitant partner type from the other data sources (the reviews of dating apps, PN intervention content and guidelines, and findings from focus group discussions). We applied the same approach to identify the key biomedical, psychological and social factors that differentiated the particular types of relationships and partners.
2. **Multidisciplinary clinical expert input:** Experts provided opinions and suggestions in the following sequence; 1) we discussed initial drafts of the matrix with the full LUSTRUM project team, which includes clinical sexual health and PN specialists, 2) we sought opinions on a revised draft from multidisciplinary clinical sexual health care professionals in a workshop on PN outcomes at a national specialist conference (BASHH Annual Spring Meeting, 2017), 3) a senior team member with clinical expertise (CSE) applied the feedback from the workshop participants to examine the conceptual coherence and logic of the matrix. She assessed its utility against a range of real and hypothetical patient scenarios and discussed areas of uncertainty and disagreement with the LUSTRUM team, 4) we simplified the matrix again to improve clinical utility. This process produced the sex partner classification that the project team considered clinically useful, 5) STI clinical, public health and epidemiology experts from UK, Australia and The Netherlands gave further input as part of a BASHH working group to develop PN outcomes in May 2019, and changes were incorporated.
3. **Piloting the classification:** We developed a standardised 15-minute training session for healthcare professionals about the refined classification and how to use it during sexual history-taking and PN consultations. The training was part of a randomised controlled trial (RCT) of accelerated partner therapy (APT) for PN for heterosexual people with chlamydia.^23^ Around 120 healthcare professionals (nurses, health advisers and doctors) received the training between 31/05/2018 and 26/09/2018 at 17 sexual health clinics in England and Scotland, which were purposively selected for their contrasting patient populations and geographical contexts. At the end of the training, we administered an informal quiz using patient scenarios. Healthcare professionals practised using the classification and discussed answers collectively. Healthcare professionals used the classification during the pre-trial baseline data collection phase and the first trial period (04/11/2018 to 28/04/2019). As part of the trial’s monthly clinic support sessions, we asked each clinic’s “trial champion” about their team’s experiences using the classification for sexual histories and PN, specifically noting any instances where clinicians had felt unable to assign a particular patient’s partner to any category.
4. **Applying the Theoretical Domains Framework (TDF) to the classification:** To understand how the classification might enhance PN, we explored how, from an index patient’s perspective, relative barriers and facilitators to engaging with PN differed according to partnership type. We used the TDF to code these findings.^19^ In this way, we outlined the differential barriers and facilitators shaping index patient engagement with PN in order to speculate about how the classification might suggest particular tailored PN approaches for different partner types.

## Results

The results are presented in relation to the research questions.

### 1. Which labels are used to classify sexual partners and which biomedical, psychological and social aspects differentiate them?

The evidence integration initially resulted in eight categories, which summarised the range of identified sexual partner types (Figure 1, top row). These types span (from left to right) the traditional dichotomy from “regular” to “casual”. The partner types also incorporate trajectories across time, with intensity and duration decreasing from left to right. We identified eight important variables: two in the biomedical domain, four psychological factors and two social aspects, which could help distinguish between partnership types.

**Figure 1:**
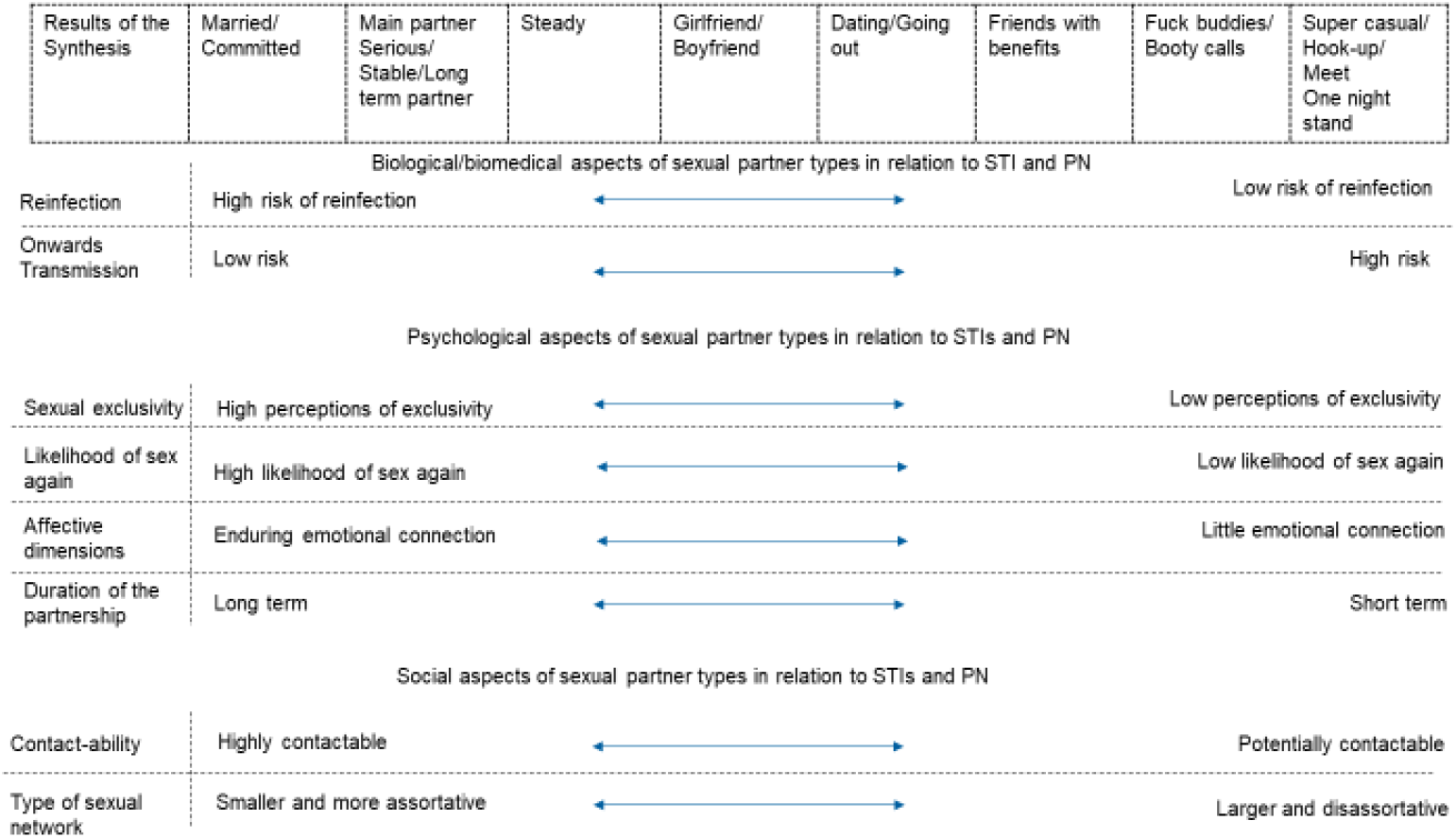
Initial matrix of sexual partner types and their relation to key factors that differentiate them. Evidence synthesis derived an initial eight partner types (top row). Factors which differentiated them are shown in the left hand column.

From a biomedical perspective, the classification captures the concept that both the likelihood of STI reinfection within the partnership and onwards transmission to other people differ by partner type.^1^ Reinfection within a partnership is more pertinent for partner types such as ‘married/committed’ and ‘main partner’/‘serious partner’/‘stable partner’/‘long term partner’, whilst onward transmission is more of a risk at the opposite end of the partner spectrum (‘super casual’/‘hook up’/‘meet’ and ‘one-night stand’).

We identified four psychological factors that appeared to differ between partner types; higher perceptions of an exclusive partnership, higher likelihood of sex again and more enduring emotional connection are associated with those types at the left-hand side (i.e., towards ‘married’/‘committed’). In contrast, types characterised by lower expectations of exclusivity, lower likelihood of sexual intercourse with that partner again, little emotional connection and shorter duration cluster on the right-hand side towards the ‘super casual’/‘hook-up’/‘meet’/‘one-night stand’ partner types.

Social factors also distinguish between partner types. For example, partner types towards the right-hand side of Figure 1 are more likely to be embedded within larger, disassortative, multifaceted sexual networks than those towards the left-hand side. Contactability is less clearly polarised and may be possible all across the partner spectrum but is likely to reduce from left to right.

### 2. What does a classification of sexual partners look like for clinical practice?

We simplified the initial classification from eight to four categories to make it practical for use in clinical care, based on the multidisciplinary clinical expert input (Figure 2). The clinician researchers recommended adding a fifth category of sex partner “Sex worker”, which had not emerged from the scoping review as a separate partnership type. A separate category for sex work reflects UK national guidance on sexual history-taking ^24^ and because PN and risk reduction strategies are likely to differ for this group. The research team assigned short neutral labels to each category as an aide-memoire for health care professionals. The labels are not intended to be used as descriptors in consultations with patients although some of the words used may figure in patients’ descriptions of their relationships, e.g. “One-offs”, “Sex worker”.

**Figure 2:**
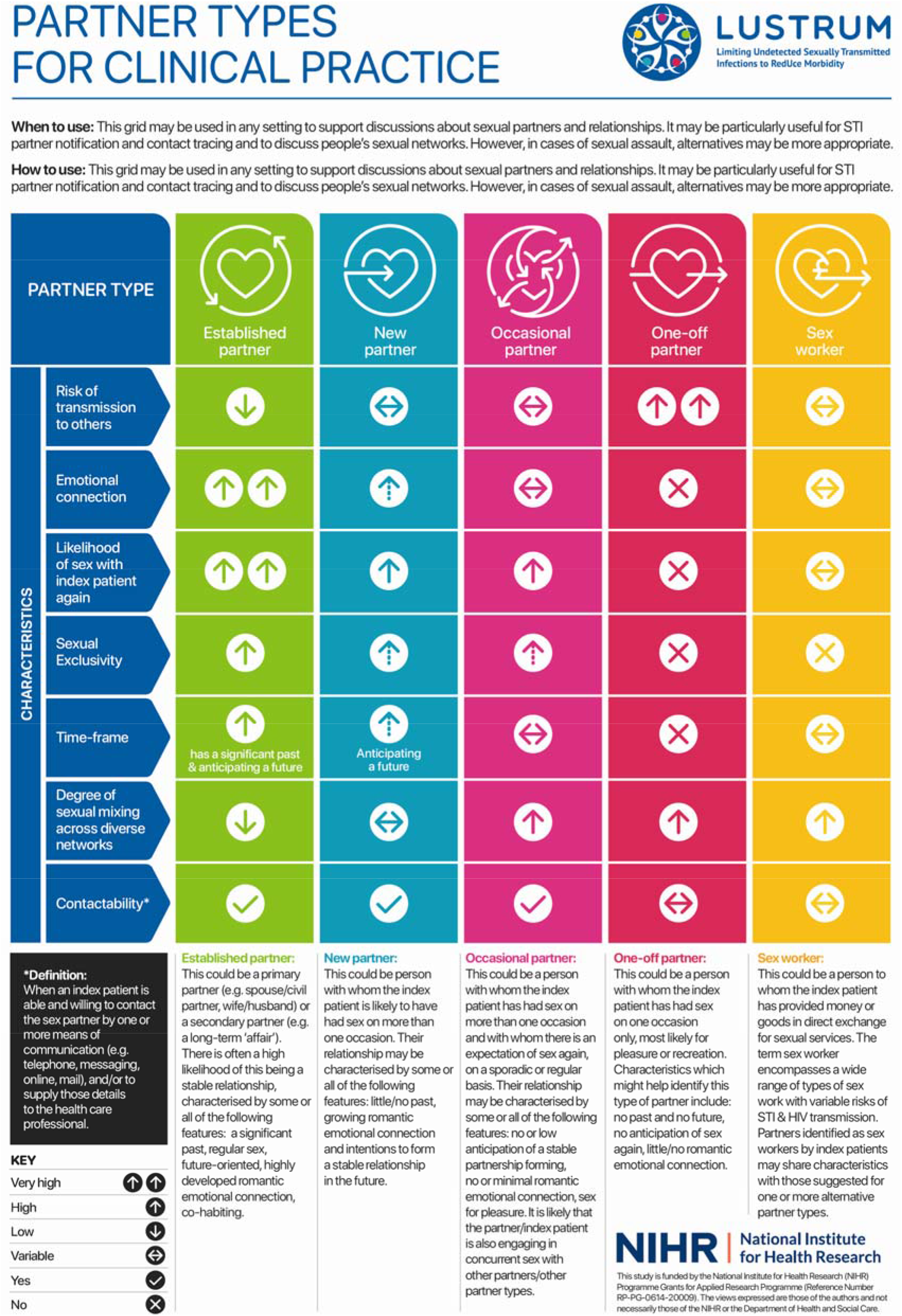
Partner Types for clinical practice.

### 3. Is the classification acceptable and feasible for use in routine clinical practice?

Informal verbal feedback from the pre-trial teaching sessions was overwhelmingly positive. Participants agreed that the new categories could help overcome the well-recognised limitations of the “regular/casual” dichotomy. Participants correctly assigned partners to categories in the post-training skills test. During the trial baseline data collection phase, we discussed and resolved categories for a small number of clinical cases, raised by clinical staff. There were no further queries after starting the trial and by the end of the first trial period (04/11/2018 to 28/04/2019), clinicians across the 17 study sites had used the classification to categorise 3288 sex partners from 2223 index patients. There were no instances in which clinicians felt unable to assign a sex partner to any category. Figure 2 summarises the partner types for use by healthcare professionals in sexual health clinics.

### 4. How could use of the classification help to improve individual and public health for PN?

Table 1 illustrates the ways that the different partner types within the classification may need different PN approaches.

**Table 1:**
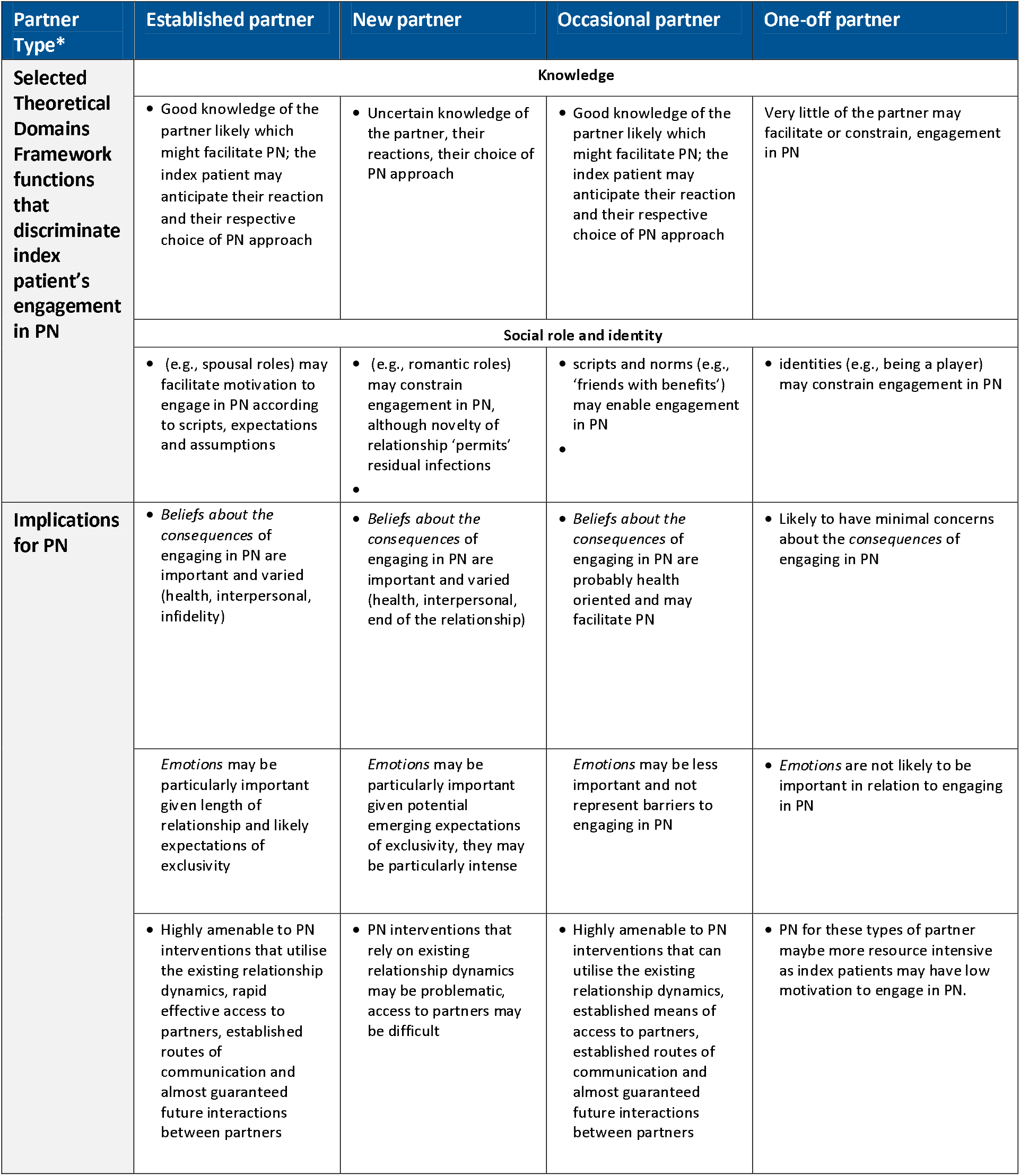
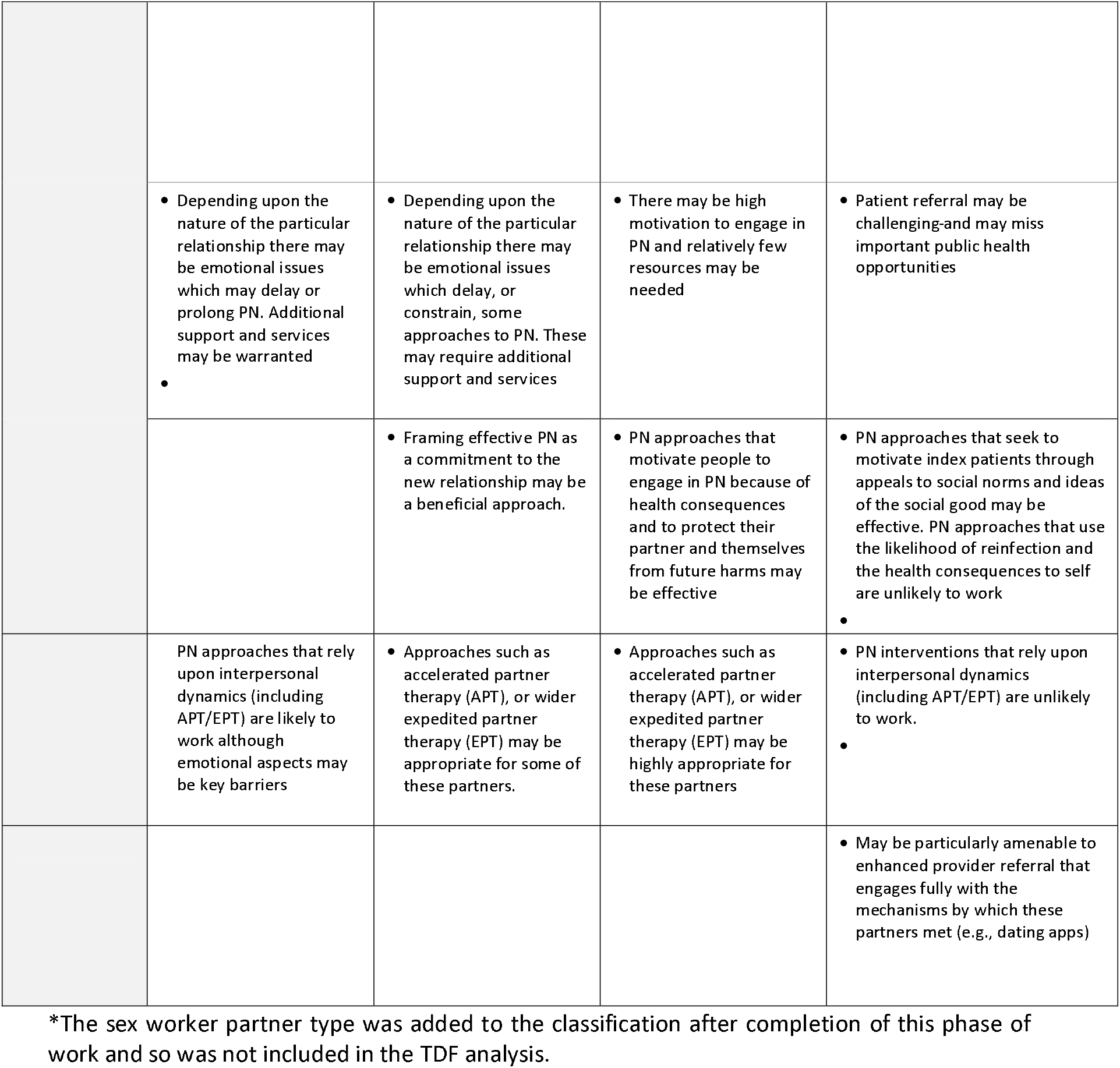
Use of Theoretical Domains Framework: Implications of the classification of partner types for partner notification approaches.

For any index patient with multiple sexual partners, the TDF^19^ suggests it may be worth exploring which type(s) of sexual partner(s) they have, and subsequently which type(s) of PN may be most appropriate for each different partner, depending on their type. Critically, although an index patient may have equal physical capability to engage in PN with diverse types of partner, there are important differences in the index patient’s motivation to engage in PN with different partner types. For example, from an index patient’s perspective, there may be very little motivation to engage in PN with ‘One-off partners’ and far more to engage with ‘Established partners’ with whom sex is likely to occur again.

For ‘Established partners’, PN approaches should utilise the deep emotional connection between partners, the likelihood of co-habitation and the high potential for reinfection. Approaches such as Expedited Partner Therapy,^25^ and Accelerated Partner Therapy^23^ are likely to be acceptable and effective. Depending on the particular relationships and expectations concerning exclusivity, PN may be taking place against a background of high emotions and potential distress; partner delivered testing/treatment options may be very useful.

For ‘New partners’, PN approaches should harness the developing emotions within such relationships and capitalise upon the relationship’s short duration. For these relationships a diagnosis of an STI can ‘make or break’ the developing relationship. For example, it may be that the STI has arisen from sex pre-dating the current ‘new’ relationship, or that the STI has been transmitted before agreements concerning exclusivity have been made.

For ‘Occasional partners’ characterised by high likelihood of the relationship having a future and likely sex again, yet limited emotions, approaches such as Expedited Partner Therapy,^25^ and Accelerated Partner Therapy^23^ may be highly acceptable.

For ‘One-off partners’, PN approaches which require an emotional connection between partners, or those that use risks of reinfection to motivate partners or are unlikely to be effective. However, given changes in the ways people meet and the widespread use of social media, index patients may well have some means of contacting these types of sexual partner. Provider referral, in which the health care professional contacts the sex partner without revealing the identity of the index patient may be useful.

## Discussion

We developed a new five-category classification of sex partner types. We synthesised diverse sources of evidence to understand the biomedical, psychological and social aspects that make the partner types identified distinct. The classification was feasible and acceptable to a range of healthcare professionals within sexual health services across England and Scotland. The classification accommodates most sex partner types described by people attending UK sexual health services and staff were able to assign all sex partners described to a category.

For use in routine clinical care, a classification needs to be pragmatic, such that the majority of partner types described in contemporary life and clinical practice can be assigned to a reasonable number of categories, whilst recognising that some patients will describe partners who cannot be neatly assigned to any category. Our proposed classification goes beyond the mutually exclusive dichotomy of ‘regular’or ‘casual’ partnerships that has been used in sexual health practice to date. By synthesising diverse sources of evidence, our classification considers the realities and increasing complexities of the contemporary social organisation of sexual relationships.

This work has drawn on, and further developed, existing classifications that typically focus on a single dataset and/or consider fewer partnership-specific variables to differentiate between the types identified. The classification has important differences from an earlier classification that was based on responses to questions in the third British National Survey of Sexual Attitudes and Lifestyles (Natsal-3).^16^ Questions in Natsal-3 distinguished between partnerships that involved cohabiting and those that were considered as ‘now steady’. We now propose using the collective label of ‘Established’. Whilst the earlier classification had just one category for ‘casual’ partners, we now propose two categories: ‘Occasional’ partners and ‘One-off’ partners. This distinction is helpful because of the greater heterogeneity in casual sex (and the labels attributed to this) as compared with more established partnerships. The distinction is also relevant to the delivery of PN, reflecting variation in the extent that different types of casual partners can be traced and/or contacted.

The partner types that emerged are culturally embedded in UK sexual health settings. Although we searched the international literature, the classification might not be generalisable to very different populations or cultures. Whilst we piloted the usability of the classification during a trial that included only people who have sex with opposite gender partners, the partner types also make sense for same sex partnerships and those that include trans/transgender and non-binary people. The classification takes account of current societal sexual behaviours and so may not be relevant if significant shifts in sexual behaviours occur.

A pragmatic, evidence-informed classification could enhance clinical practice and research study design. More appropriate targeting and tailoring of PN and other sexual health interventions should result in greater individual and public health benefits. Whilst our classification prioritises utility within the clinical context rather than the general population, it is informed by published evidence and primary research undertaken with people in a variety of settings, including clinic attendees and lay people.

The classification could improve the ability of services to address the aims of PN at both individual patient level (prevention of reinfection) and public health level (transmission prevention) by ensuring that the best available evidence guides the choice of PN methods offered by services. Tailored PN approaches should enable more effective targeting of resources and audit that is meaningful in epidemiological terms, as well as relating to the individual index patient. This methodological advance will also enhance social epidemiology and the evidence provided by behavioural surveillance to facilitate development of patient-centred risk assessment tools. Such tools will enable robust comparisons of the transmission prevention outcomes of existing and novel PN approaches as well as index patient-centred outcomes. Collectively, these advances could improve patient care by ensuring that best-available evidence guides choice of PN methods offered by services. At the individual patient level, an awareness of the distinct aspects of each partner type could enable better tailoring of PN interventions offered by health care professionals and allow a more strategic approach to prevention of transmission. This offers considerable potential when PN is particularly important at both the individual and public health level, such as with cases of extensively drug-resistant pathogens.^26^

The content validity of the classification is being evaluated in the RCT of accelerated partner therapy,^23^ which will include analysis of trial outcomes by partner type. Evaluation in clinical practice through a UK national audit will take place in 2020 and will establish whether the classification accommodates most sex partner descriptions, including same sex partners, when embedded in routine care. Additional studies are needed to determine the utility of the classification to improve measurement of the impact of PN strategies and help focus resources. Future work will address tailored intervention development based on partner type, which could inform targeting of resources to reach sex partners who might contribute disproportionately to transmission within the population. New PN methods will need to embrace the range of communication technologies used within contemporary social and sexual networks and determine the cost-effectiveness of PN approaches with different types of partner in relation to reducing onwards transmission at the population level.

## Data Availability

Data are available from the corresponding author upon reasonable request

## Supplementary File

**Table S1:**
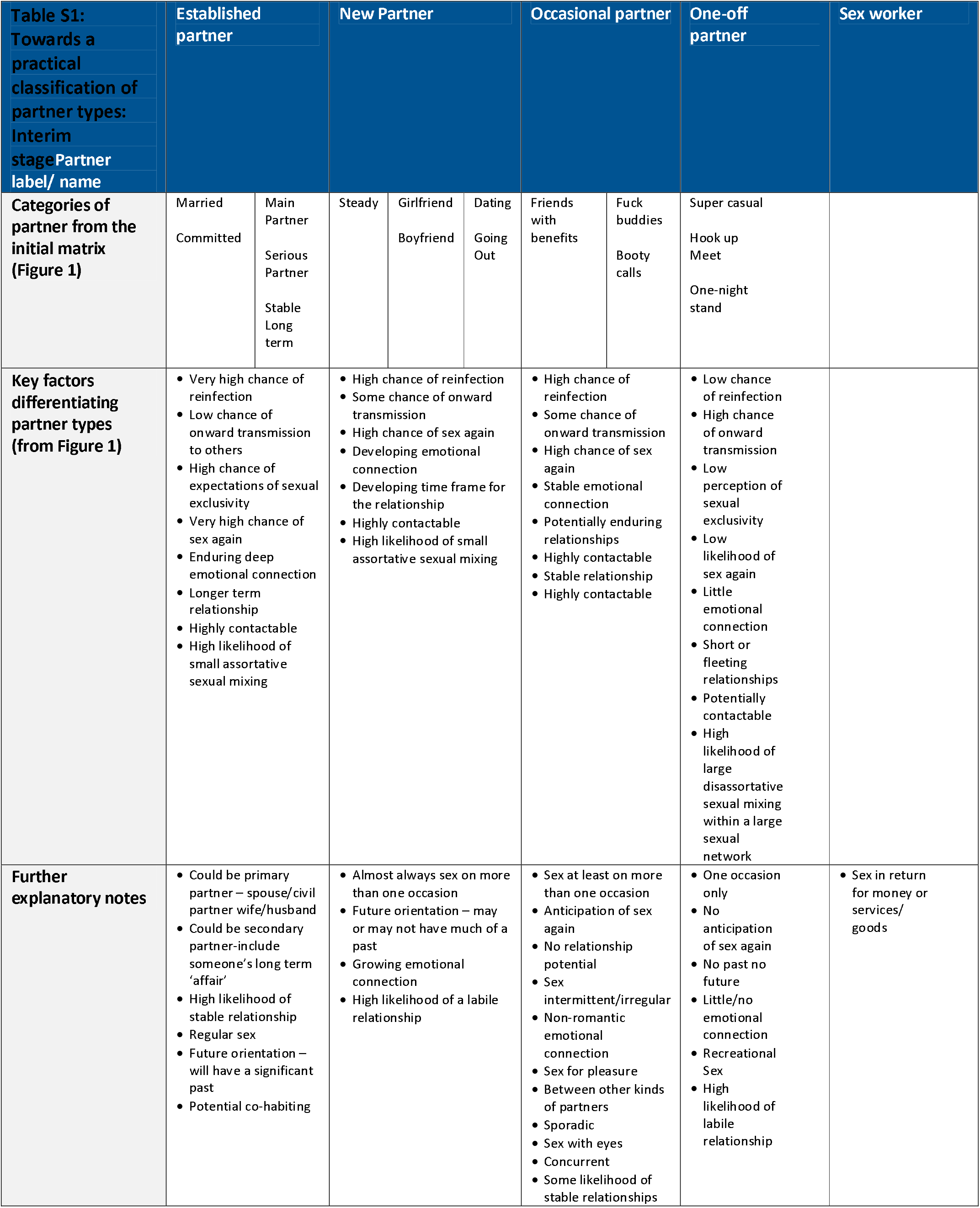
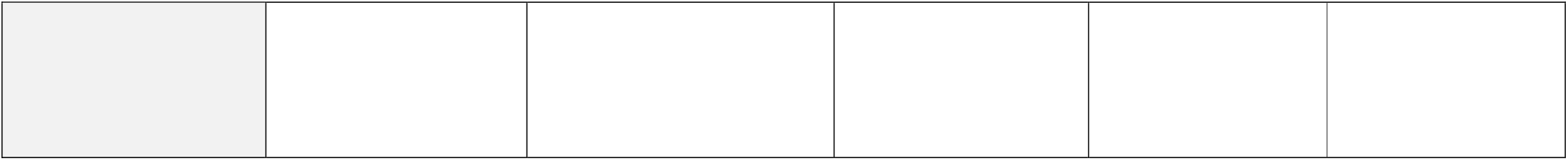
The first row of the table shows the condensed, five-type partner typology, with the second row illustrating how they relate to the ‘original’ eight partner types described in Figure 1. The third row summarises the spectrum of issues detailed in Figure 1.

## Acknowledgements

We would like to thank all of the participants who took part in focus groups and interviews and all individuals and organisations who helped with recruitment for this study via social media, word of mouth and direct contact to their networks. We are also very grateful to the expert group who attended the BASHH PN Outcomes workshop (Ann Sullivan, Ana Harb, Martin Murchie, Ceri Evans, Jonathon O’Sullivan, Hannelore Gotz, Jane Hocking). Thanks to Ceri Evans and Sophie Herbert for their input into Figure 2 and to Lock Cheung Hwa at Diva Creative for the graphic design; thanks to Morgan Williamson for help with figure 1. This study has been shaped through ongoing discussion and support from the whole LUSTRUM team (Claudia S Estcourt (Principal Investigator), Alex Comer, Alison R Howarth, Andrew Copas, Anna Tostevin, Anne M Johnson, Catherine H Mercer, Chidubem (Duby) Ogwulu, Christian Althaus, Fiona Mapp, Gabriele Vojt, Jackie Cassell, Jean McQueen, John Saunders, Karen Pickering, Maria Pothoulaki, Melvina Woode Owusu, Merle Symonds, Nicola Low, Oliver Stirrup, Paul Flowers, Rak Nandwani, Sonali Wayal, Susannah Brice, Tracy Roberts) and the Programme Steering Committee (PSC) (Simon Barton (chair), Alex Miners, Artemis Koukounari, David Crundwell, Emmanuel Rollings-Kamara, Lynis Lewis, Rebecca Turner, Robbie Currie, Rachel Shaw, Saima Saddiqui (on behalf of NIHR programme management).

## Ethical approval

Ethical approval received from Glasgow Caledonian University Research Ethics Committee (HLS/PSWAHS/A15/256) and NHS Ethics Approval (16/NI/0211) were obtained. Trial: Ethical approval received from London Chelsea Research Ethics Committee (18/LO/0773).

